# A Novel AI-based Score for Assessing the Prognostic Value of Intra-Epithelial Lymphocytes in Oral Epithelial Dysplasia

**DOI:** 10.1101/2024.03.27.24304967

**Authors:** Adam J Shephard, Hanya Mahmood, Shan E Ahmed Raza, Syed Ali Khurram, Nasir M Rajpoot

## Abstract

Oral epithelial dysplasia (OED) poses a significant clinical challenge due to its potential for malignant transformation and the lack of reliable prognostic markers. Current grading systems for OED may not be reliable for prediction of malignant transformation and suffer from considerable inter- and intra-rater variability, potentially leading to suboptimal treatment decisions. Recent studies have highlighted the potential prognostic significance of peri-epithelial lymphocytes (PELs) in malignant transformation, with suggestions that intra-epithelial lymphocytes (IELs) may also play a role. In this study, we propose a novel artificial intelligence (AI) based IEL score from Haematoxylin and Eosin (H&E) stained Whole Slide Images (WSIs) of OED tissue slides. We further determine the prognostic value of our IEL score on a large digital dataset of 219 OED WSIs (acquired using three different scanners), compared to pathologist-led clinical grading. Notably, despite IELs not being incorporated into the current WHO grading system for OED, our findings suggest that IEL scores carry significant prognostic value that were shown to further improve both the Binary/WHO grading systems in multivariate analyses. This underscores the potential importance of IELs, and by extension our IEL score, as prognostic indicators in OED. Further validation through prospective multi-centric studies is warranted to confirm the clinical utility of the proposed IEL score and its integration into existing grading systems for OED.

**Conflict of Interest Statements:**

1. NMR is the co-founder, CEO and CSO of Histofy Ltd., UK. He is also the GSK Chair of Computational Pathology and is in receipt of research funding from GSK and AstraZeneca.
2. SAK is a shareholder of Histofy Ltd.
3. All other authors have no competing interests to declare.

## 1. Introduction

Head and neck cancer encompasses a diverse group of malignancies originating from the upper aerodigestive tract, including the oral cavity, nasal cavity, pharynx, larynx, salivary glands and sinuses [1]. Among these, oral squamous cell carcinoma (OSCC) stands as one of the most prevalent subtypes, predominantly affecting the oral mucosa and accounting for a significant proportion of head and neck cancer cases [1]. OSCC arises from the squamous epithelial cells lining the oral cavity and is strongly associated with risk factors such as tobacco use, alcohol consumption, and human papillomavirus (HPV) infection [2].

Characterised by aggressive local invasion and potential for regional and distant metastasis, OSCC poses considerable challenges in diagnosis and management. Combination therapy approaches including surgery, radiation therapy, and chemotherapy are often employed [3], which even if successful, are associated with functional problems including masticatory, speech and swallowing impairments, drastically affecting quality of life [4]. Prognosis for advanced stage OSCC is poor, having a five-year survival rate of just 40% [5]. This drastically increases with early diagnosis to 80-90% [5], highlighting the huge benefits of early detection.

Oral cancer lesions are typically preceded by a pre-cancerous state, a group of lesions termed oral potentially malignant disorders (OPMDs) including homogeneous/non-homogeneous leucoplakia (white/white-red speckled lesions) or erythroplakia (red lesions) [2,6]. Biopsies of the lesions enable microscopic examination by histopathologists to determine the presence or absence of pre-cancer (oral epithelial dysplasia or OED) or cancer. Lesions of the oral mucosa exhibiting dysplasia are statistically more likely to transition into OSCC than non-dysplastic lesions [7].

The histopathological grading of Haematoxylin and Eosin (H&E) stained tissue using the World Health Organisation (WHO, 2017 [8]) classification system remains the current accepted practice for diagnosis and risk stratification of OED lesions. This system categorises OED into three grades: mild, moderate, and severe, based on the presence, severity, and location of various cytological and architectural histological features (28 in total [9,10]). However, this approach has been widely criticised for its significant intra- and inter-observer variability and its limited predictive ability for malignant transformation risk, which can impact patient management. An alternate binary grading system has been proposed to address the WHO grading shortcomings, and to improve the reproducibility of grading [11,12]. This system classifies lesions as either low- or high-risk based on the number of cytological and architectural features outlined in the WHO criteria. Mahmood *et al.* [13] showed the utlity of various of these pathologist-assigned histological features in prognostic models. However, overall, studies have shown that both the three-tier and binary grading systems suffer from significant variability and unreliability [14,15]. This underscores the need for more objective and reproducible methods and *features* (markers) for grading OED that can better predict the risk of malignant transformation.

Recent advancements in digital pathology have facilitated the digitisation of histology slides into whole slide images (WSIs) through high-resolution digital scanners. This has spurred significant growth in computational pathology [16,17]. Concurrently, the evolution of new deep learning techniques has complemented both pathology and radiology, enabling the automation of pipelines and demonstrating the potential of deep learning in predicting patient outcomes [16–18]. In the emerging area of computational pathology, deep learning has been applied to automatically segment epithelium across various histology images (e.g., oral, cervical, prostate) [19–23] and to further segment and classify individual nuclei within WSIs [24,25]. In the context of OED, our previous work has used deep learning to segment dysplasia [19] and also the oral epithelium into sub-regions: the lower basal layer, the middle epithelial layer, and the superior keratin layer [20,21]. These methods have even been used to predict OED malignant transformation, based on either deep [26] or nuclear features [20,27]. Thus, deep learning tools offer a potential avenue for reducing grading variability while ensuring consistency across sites in informing treatment decisions [28,29].

Computational pathology has not only allowed researchers to replicate and automate pathology workflows, but also to aid in biomarker discovery. Bashir *et al.* [26] developed a pipeline to predict malignant transformation in OED. In doing so they discovered a positive association between an increased presence of peri-epithelial lymphocytes (PELs) and transformation. Further, Shephard *et al.’s* [20] work also suggested the potential association between both PELs and intra-epithelial lymphocytes (IELs) and OED malignant transformation. These studies highlight the need for further exploration and validation of the role of IELs in OED.

IELs are small, round mononuclear white blood cell lymphocytes found within the epithelial layer in the paracellular space between epithelial cells [30]. They are found in the skin and within the epithelial layer lining the intestine, lungs, reproductive tract and oral cavity, and are typically thought to be T lymphocytes [30,31]. In the gastrointestinal (GI) tract, they are components of gut-associated lymphoid tissue. Within normal mice, there is one IEL per 5-10 epithelial cells in the small intestine. In human duodenal biopsies, healthy individuals usually have less than 5-10 IELs per 100 epithelial cells; however, this number can increase significantly, and is a hallmark of coeliac disease [31]. In the oral cavity, the inflammatory response and the role of IELs is poorly understood [32]. As yet, neither PELs nor IELs are used as markers within the WHO or binary grading system.

While the role of lymphocytes in cancer immunity is well-documented, their significance in dysplasia remains underexplored. Therefore, we present an in-depth exploration of IELs within OED, and investigate the prognostic value of a digital IEL score. This approach aligns with other automated methods within the computational pathology community, such as Tumour-Infiltrating Lymphocyte (TIL) scoring [33] and Mitosis Counting (MC) [34]. We aim to elucidate the prognostic utility of AI-generated IEL scores in OED, utilising advanced image analysis techniques to quantify lymphocytic infiltration within the epithelium. Through comprehensive evaluation of the correlation between IEL scores and clinical outcomes using a relatively large dataset (in the context of OED), we explore the potential of the proposed IEL score as a prognostic indicator. In the spirit of reproducibility, we also release the full inference pipeline for generating our AI-generated IEL scores, based on H&E-stained WSIs from oral tissue sections, <u>adamshephard/oed_iel_scoring (github.com)</u>.

## 2. Materials and Methods

### 2.1 Study Data

The study data consisted of a retrospective cohort of histology tissue sections (dating 2008 to 2016 with minimum five-year follow-up data) collected from the Oral and Maxillofacial Pathology archive at the School of Clinical Dentistry, University of Sheffield, UK. After microscopic inspection of the tissue sections by a Consultant Pathologist (SAK), newly cut 4 µm sections of the selected cases were obtained from formalin fixed paraffin embedded blocks and stained with H&E for analysis. Ethical approval was obtained by the NHS Health Research Authority West Midlands (Ref: 18/WM/0335) and experiments were conducted in compliance with the Declaration of Helsinki.

In total, 219 slides were collected from 188 patients. The slides were digitised to high-resolution WSIs at 40× objective power using one of three scanners: NanoZoomer S360 (Hamamatsu Photonics, Japan; 0.2258 mpp), Aperio CS2 (Leica Biosystems, Germany; 0.2520 mpp), Pannoramic 1000 (P1000, 3DHISTECH Ltd, Hungary; 0.2426 mpp). Clinical data for the cohort included patient age (at time of diagnosis), sex, intraoral site, OED grade (using binary and WHO 2017 systems) and transformation status. Transformation was defined as the progression of a dysplastic lesion to OSCC at the same clinical site within the follow-up period, and time to transformation was measured in months. To ensure diagnostic consistency, all cases were evaluated by at least two certified pathologists (PMS, PMF, KH, DJB), who provided an initial diagnosis based on the WHO grading system (between 2008-2016). To confirm the WHO (2017) grade and assign binary grades, the cases were blindly re-evaluated by SAK and a clinician with a specialist interest and expertise in OED analysis (HM). In total, 42 patients (49 WSIs) developed malignant transformation. An overview of the dataset is given in **Table 1**.

**Table 1.**
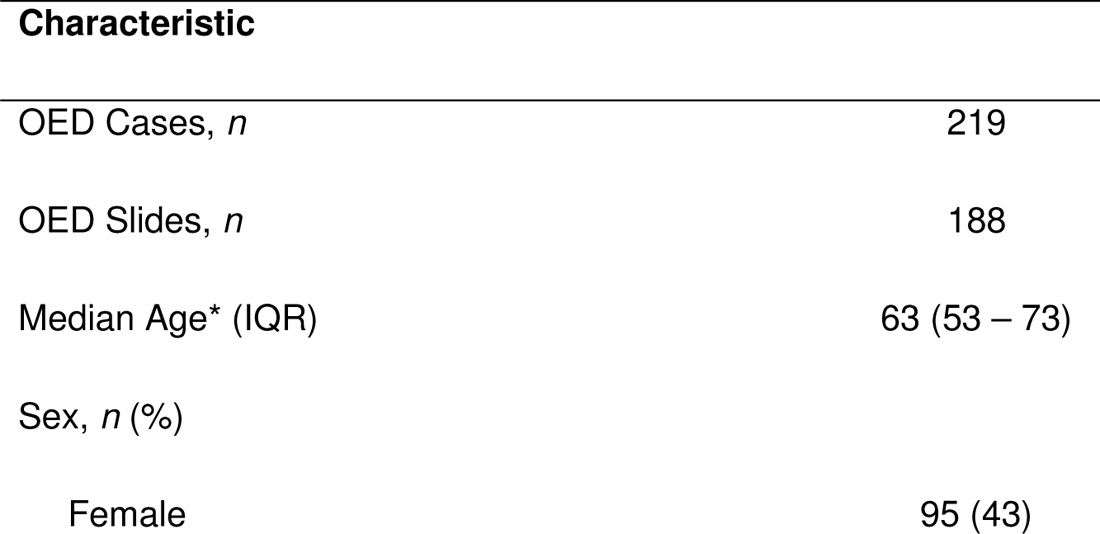
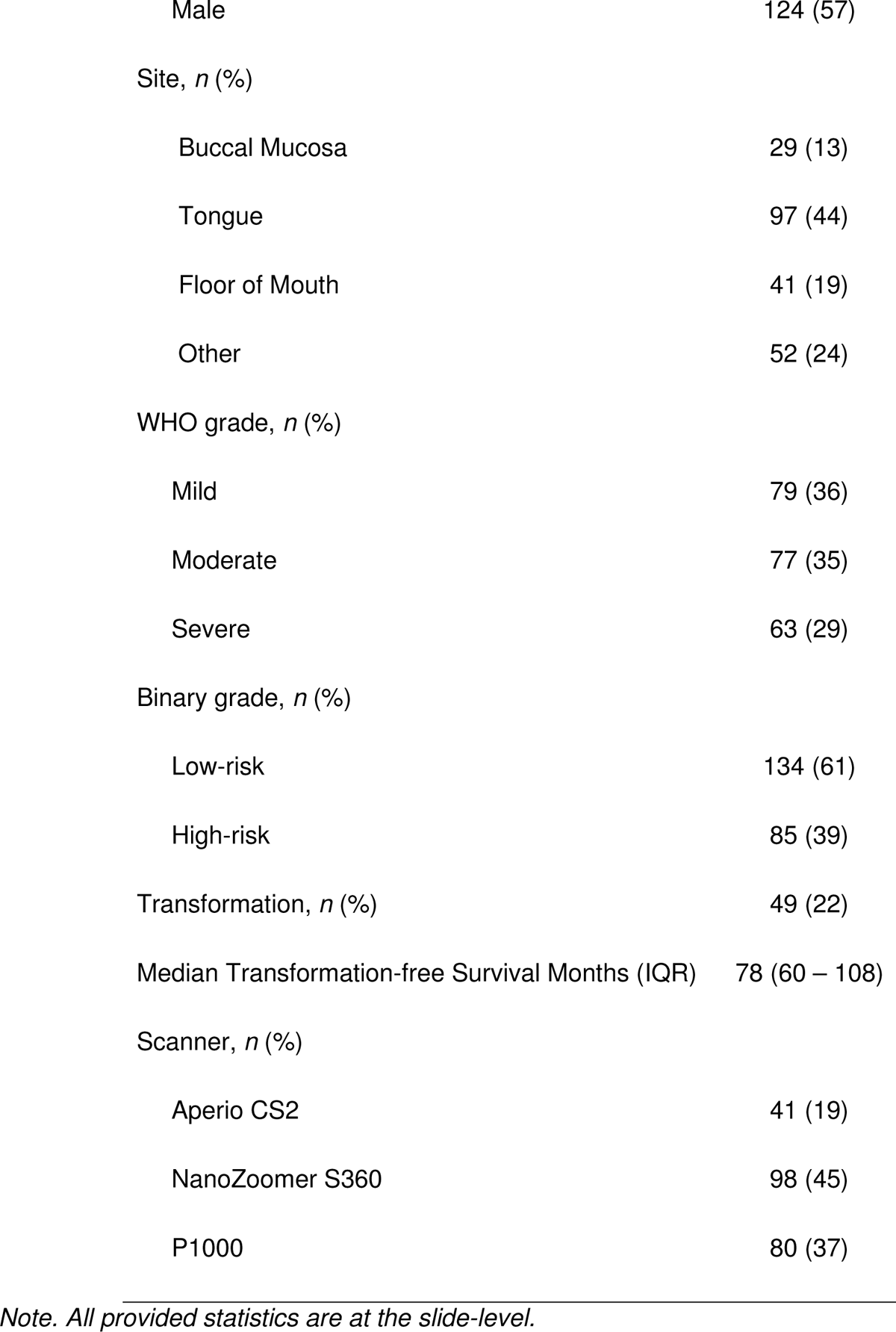
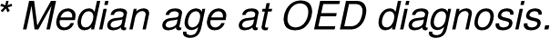
Overview of OED samples included in this study.

| Characteristic |  |
| --- | --- |
| OED Cases, *n* | 219 |
| OED Slides, *n* | 188 |
| Median Age* (IQR) | 63 (53 – 73) |
| Sex, *n* (%) |  |
| Female | 95 (43) |
| Male | 124 (57) |
| Site, *n* (%) |  |
| Buccal Mucosa | 29 (13) |
| Tongue | 97 (44) |
| Floor of Mouth | 41 (19) |
| Other | 52 (24) |
| WHO grade, *n* (%) |  |
| Mild | 79 (36) |
| Moderate | 77 (35) |
| Severe | 63 (29) |
| Binary grade, *n* (%) |  |
| Low-risk | 134 (61) |
| High-risk | 85 (39) |
| Transformation, *n* (%) | 49 (22) |
| Median Transformation-free Survival Months (IQR) | 78 (60 – 108) |
| Scanner, *n* (%) |  |
| Aperio CS2 | 41 (19) |
| NanoZoomer S360 | 98 (45) |
| P1000 | 80 (37) |

*Note. All provided statistics are at the slide-level.*
\* Median age at OED diagnosis.

### 2.2 Deep Learning Framework

#### 2.2.1 Dysplasia and Nuclear Segmentation

Since dysplastic changes may not be widespread across the entire tissue section in a slide, the first step of the AI pipeline involved identification and localisation of the dysplastic tissue regions for semantic segmentation. To achieve this, we used a pretrained Transformer [19] (based on Trans-UNet [35]) that automatically detects and segments the different dysplastic regions in a H&E-stained oral tissue WSIs. Further, an additional pretrained CNN-based HoVer-Net+ model [20,21] was used to segment the epithelium and the individual nuclei across each WSI. This model classifies nuclei within the epithelium as being “epithelial” or “other” nuclei. In OED tissue, aside from epithelial nuclei, only lymphocytes are expected to be in the epithelium. Thus, we classify these “other” nuclei as IELs.

#### 2.2.2 Intra-Epithelial Lymphocyte (IEL) Scoring

For IEL scoring, we counted the number of IELs within the dysplasia regions alone, and we used these counts to generate the following IEL scores:

1. The IEL Index (II) – the **number of IELs** per unit area of dysplasia, within the **entire** dysplastic region of the WSI
2. The IEL Peak Index (IPI) – the **maximum number of IELs** per unit area of dysplasia in **any given area** of dysplasia (here, chosen to be a patch of size 512 x 512, at 1.0 mpp resolution)
3. The IEL Count (IC) – the **number of IELs per 100 dysplastic epithelial cells**, in **any given area** of dysplasia (here, chosen to be a patch of size 512 x 512, at 1.0 mpp resolution)
4. The IEL Peak Count (IPC) – the **maximum number of IELs per 100 dysplastic epithelial cells**, within the **entire** dysplastic region of the WSI In **Figure 1**, we provide an overview of the proposed analytical pipeline used to generate our IEL scores.

**Figure 1.**
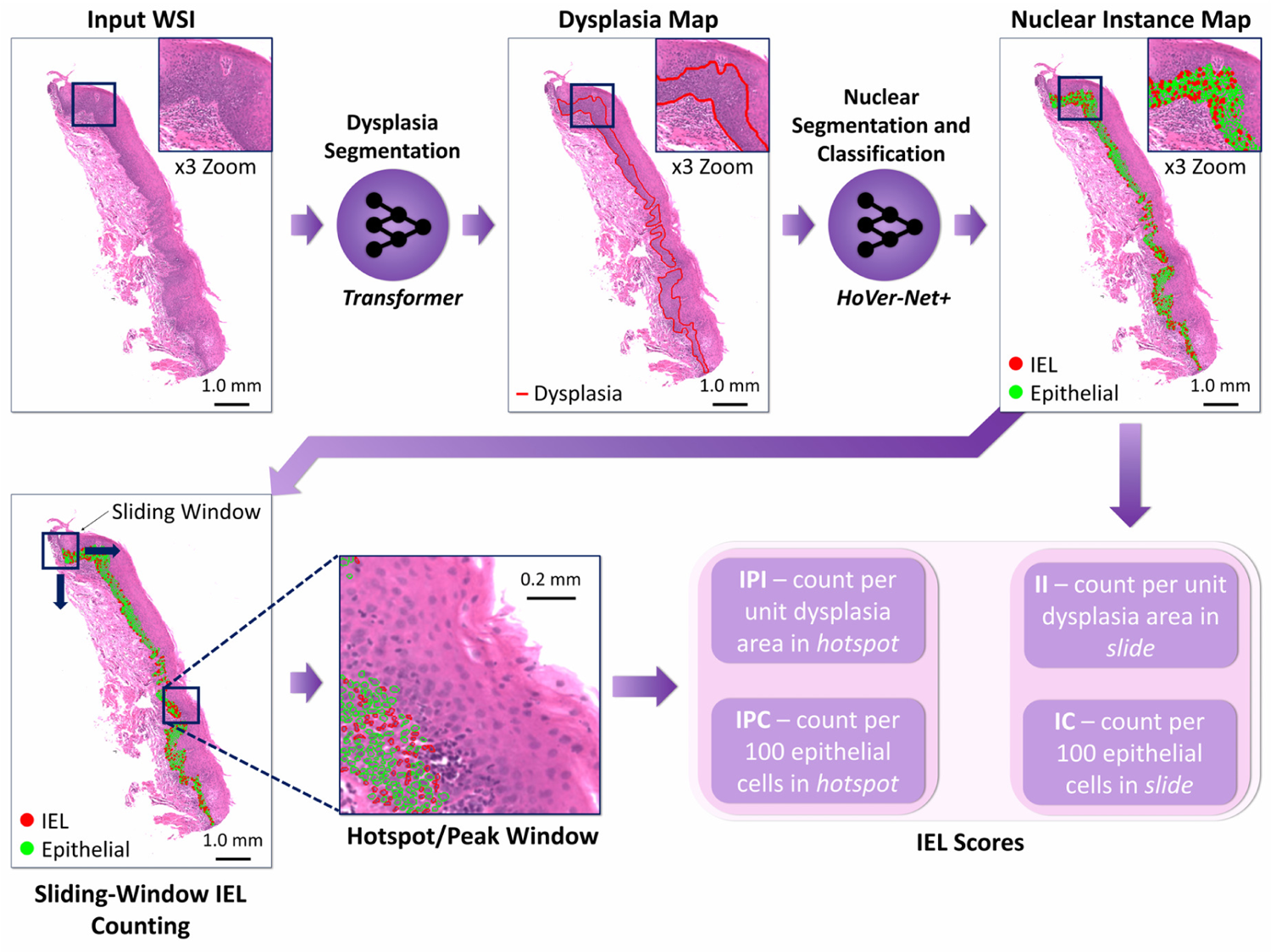
Overview of the pipeline used to generate IEL scores. An input WSI first goes through the Trans-UNet model for dysplasia segmentation. Following this, we perform nuclear segmentation using HoVer-Net+. We then generate the II and IC scores based on the IELs and epithelial nuclei detected in the dysplastic regions. For the IPL and IPC scores we find the window with the highest value for that score, using a sliding window approach.

### 2.3 Statistical Analysis

Survival analyses were conducted to assess the prognostic significance of the IEL scores in predicting transformation-free survival. Cases were split into low- and high-risk groups based on whether their IEL score was lesser/greater than the mean IEL score. We used the mean IEL score, owing to the imbalance in the number of cases transforming to malignancy (22%), and therefore did not necessarily require a 50-50 split in low-/high-risk groups. Kaplan-Meier curves were generated, and long-rank tests were used to determine the statistical significance of this stratification (for IEL scores, WHO and binary grades). We used concordance index (C-index) to measure the rank correlation between the scores and patients’ survival time. A univariate Cox proportional hazards model was employed to determine the prognostic utility of the IEL scores compared to the WHO grade, binary grade, sex, age and lesion site, to predict transformation-free survival. Transformations were right censored at eight years. We, therefore, additionally used the hazard ratio (HR) and *p*-value generated from the univariate analyses as further metrics for evaluation. For reporting, we focus on the *p*-value from the proportional hazard model analyses, over that of the log-rank test, since both tests share the same assumptions, but Cox PH models allow for the use of continuous exposure variables [36]. However, for completeness we also provide the log-rank *p-*value with the Kaplan-Meier curves.

Finally, we performed post-hoc analyses to compare the AI-assigned IEL scores between cases that exhibited transformation and those that did not, using two-tailed t-tests to determine statistical significance. We calculated effect sizes for these tests (Cohen’s *d*). We additionally did these analyses for both WHO and binary grades.

## 3. Results

### 3.1 Survival Analyses

We compared the various digital methods for IEL scoring to the other grading systems through Kaplan-Meier curves (see **Figure 2**), to demonstrate their prognostic utility. The binary grading scheme showed the clearest separation between low- and high-risk cases (C-index = 0.74, *p* < 0.001). For ease of comparison, we divided the WHO grades into two groupings: WHO G1, where mild cases were compared against moderate and severe cases combined; and WHO G2, where mild and moderate cases combined were compared against severe cases. The WHO G1 grouping showed a clear separation between cases (C-index = 0.67, *p* < 0.001), whereas the WHO G2 grouping showed less clear stratification (C-index = 0.62, *p* < 0.001). We introduced four different IEL scores in this work. Cases were split into low- and high-risk groups based on the IEL score, according to the mean IEL score. Interestingly, the IEL scores generated based on the number of IELs per 100 dysplastic epithelial cells (i.e., IC and IPC; as opposed to per unit area), gave the best separation between low- and high-risk cases. Both the IC and IPC scores gained the highest C-index of 0.67 (both *p* < 0.001), out of the IEL scores, whilst the II score gained a C-index of 0.63 (*p* = 0.003), and the IPI score gained a C-index of 0.61 (*p* = 0.015).

**Figure 2.**
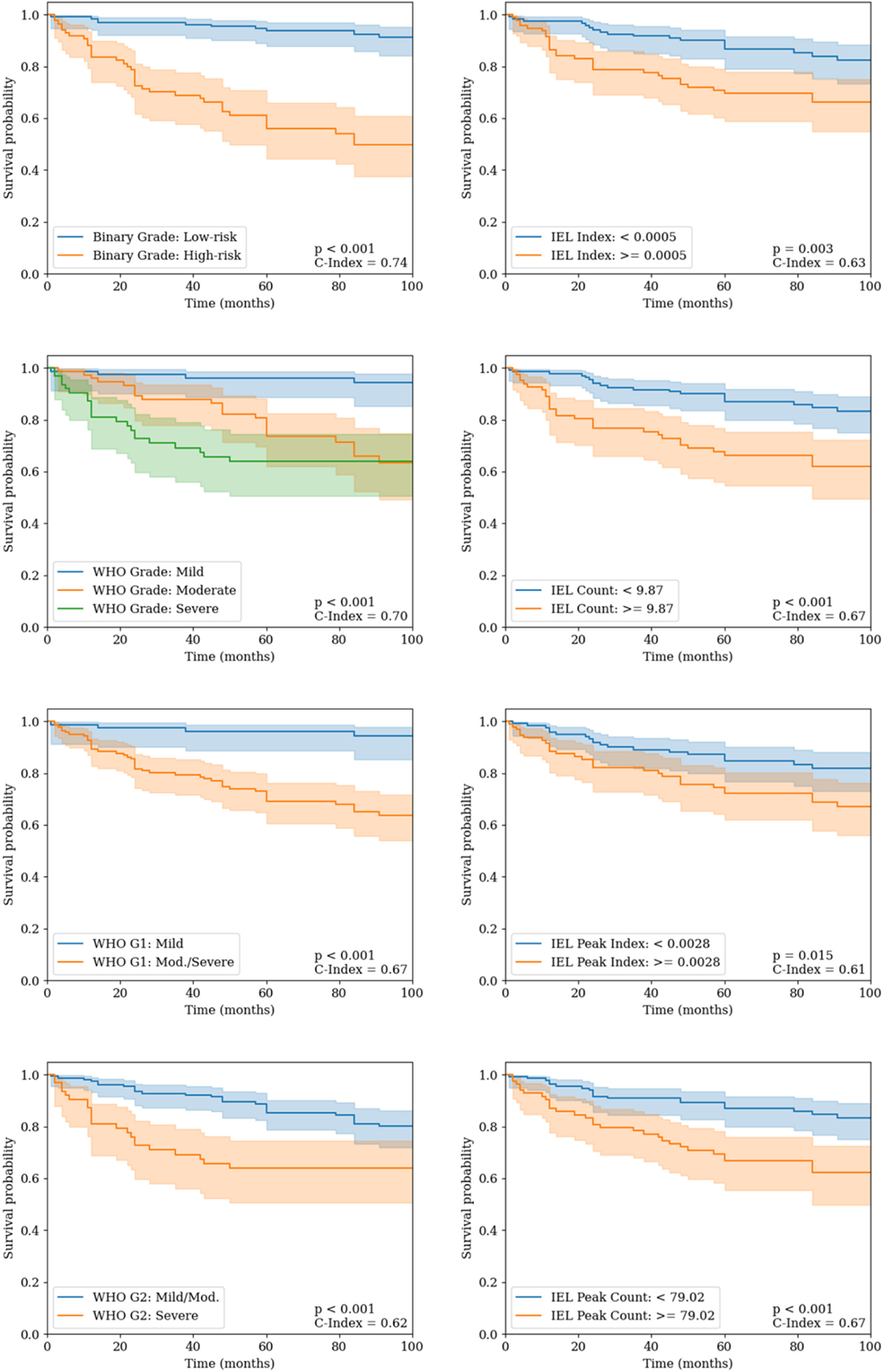
Kaplan Meier survival curves for pathologist grades (left) compared to the IEL scores (right).

See **Table 2** for the results from the univariate Cox proportional hazard models based on the clinical and digital parameters/scores. As can be observed in this table, sex, age, and lesion site appear to have no significant effect. One can also see high hazard ratios (HRs) for the binary grade and WHO grades. The HRs for the IEL scores are lower than that of the individual grades while still being somewhat significant, with IC having the highest HR = 1.65 and showing significance (*p* < 0.001). We additionally show the multivariate analyses in **Table 3**, comparing the effect of combining the above clinical and digital parameters on transformation-free survival. Within this analysis, we initially see that the IPC appeared to give some additional prognostic information (HR = 1.36, *p* = 0.003) when compared to the IC score (HR = 1.46, *p* = 0.002). Interestingly, we additionally, found that the WHO grade (HR = 0.68, *p* = 192) provided no additional prognostic utility when compared to the Binary grade (HR = 13.88, *p* < 0.001). In contrast, when we instead added the IC and IPC scores to both the Binary grade and WHO grade Cox PH models, both the C-index increased when compared to the univariate models, and the IC/IPC score were additionally shown to be significant with a high HR. Thus, these analyses demonstrate the prognostic utility of the IC and IPC scores, and the potential utility of adding IEL information to the grading system.

**Table 2.** Univariate analysis of clinical and digital parameters.

| Parameter | HR [95% CI] | *p* | C-Index |
| --- | --- | --- | --- |
| Sex | 0.97 [0.55 – 1.70] | 0.916 | 0.50 |
| Age | 1.00 [0.98 – 1.02] | 0.783 | 0.51 |
| Site | 0.97 [0.73 – 1.28] | 0.819 | 0.53 |
| WHO Grade | 2.39 [1.63 – 3.49] | **< 0.001** | 0.70 |
| WHO G1 | 8.06 [2.90 – 22.44] | **< 0.001** | 0.67 |
| WHO G2 | 2.56 [1.46 – 4.50] | **0.001** | 0.62 |
| Binary Grade | 8.20 [4.08 – 16.46] | **< 0.001** | 0.74 |
| IEL Scores |  |  |  |
| II | 1.35 [1.06 – 1.72] | **0.013** | 0.63 |
| IC | 1.65 [1.35 – 2.03] | **< 0.001** | 0.67 |
| IPI | 1.26 [0.99 – 1.61] | 0.057 | 0.61 |
| IPC | 1.52 [1.29 – 1.80] | **< 0.001** | 0.67 |

*Note.* Reported metrics are from a univariate Cox proportional hazards model. HR is the hazard ratio, where the 95% confidence interval (CI) is given in brackets. WHO G1 is mild vs moderate/severe cases. WHO G2 is mild/moderate vs severe cases.

**Table 3.** Multivariate analysis of clinical and digital parameters.

| Parameter | HR [95% CI] | *p* |
| --- | --- | --- |
| C-Index = 0.70 |  |  |
| IC | 1.46 [1.15 – 1.86] | **0.002** |
| IPC | 1.36 [1.11 – 1.68] | **0.003** |
| C-Index = 0.74 |  |  |
| Binary Grade | 13.88 [4.83 – 39.91] | **< 0.001** |
| WHO Grade | 0.68 [0.38 – 1.22] | 0.192 |
| C-Index = 0.76 |  |  |
| WHO Grade | 2.41 [1.64 – 3.55] | **< 0.001** |
| IC | 1.66 [1.34 – 2.05] | **< 0.001** |
| C-Index = 0.81 |  |  |
| Binary Grade | 9.15 [4.50 – 18.63] | **< 0.001** |
| IC | 1.88 [1.48 – 1.39] | **< 0.001** |
| C-Index = 0.76 |  |  |
| WHO Grade | 2.23 [1.52 – 3.29] | **< 0.001** |
| IPC | 1.45 [1.21 – 1.74] | **< 0.001** |
| C-Index = 0.80 |  |  |
| Binary Grade | 7.15 [3.51 – 14.55] | **< 0.001** |
| IPC | 1.30 [1.10 – 1.55] | **< 0.001** |
| C-Index = 0.81 |  |  |
| Binary Grade | 8.92 [4.24 – 18.79] | **< 0.001** |
| IC | 1.84 [1.37 – 2.48] | **< 0.001** |
| IPC | 1.03 [0.80 – 1.33] | **< 0.001** |

*Note. Reported metrics are from a multivariate Cox proportional hazards model. HR is the hazard ratio, where the 95% confidence interval (CI) is given in brackets.*

### 3.2 Post-hoc Analyses

For our post-hoc analysis, we compared the various IEL scores in cases that transformed to malignancy against those that did not (see **Figure 3**). Across all four digital IEL scores, we found significantly higher scores for cases that transformed. We also found the largest difference between groups in the IC (*d* = 0.71, *p* < 0.001) and IPC scores (*d* = 0.75, *p* < 0.001), further supporting the results of the univariate analyses. In contrast, the II and IPI scores generally had lower effect sizes, but remained significant (IC: *d* = 0.42, *p* = 0.005; and ICI: *d* = 0.33, *p* = 0.007).

**Figure 3.**
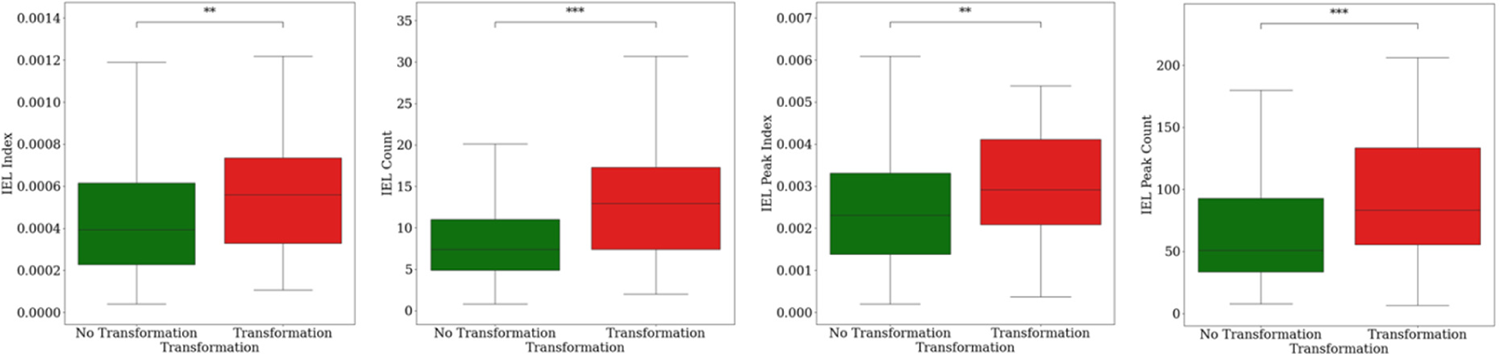
Boxplots showing the distribution of IEL scores in OED cases that did (red) and did not (green) transform to malignancy.

For the IC and IPC scores, we additionally showed how the scores varied by grade (see **Figure 4**). Our IEL scores correlate well with the binary grade, showing high-risk cases to have generally higher IC (*d* = 0.18, *p* = 0.04) and IPC scores (*d* = 0.55, *p* = 0.003). However, the effect sizes are only small-to-moderate in size, suggesting that the IEL scores are bringing new prognostic information, that isn’t necessarily incorporated by the binary grade. This further supports the multivariate analysis results. For the WHO grade, we see that more severe cases generally have higher IEL scores, however, these differences were not found to be significant for the IC score. With the IPC score, severe cases were found to have significantly higher IPC scores than mild cases (*d* = 0.59, *p* = 0.008).

**Figure 4.**
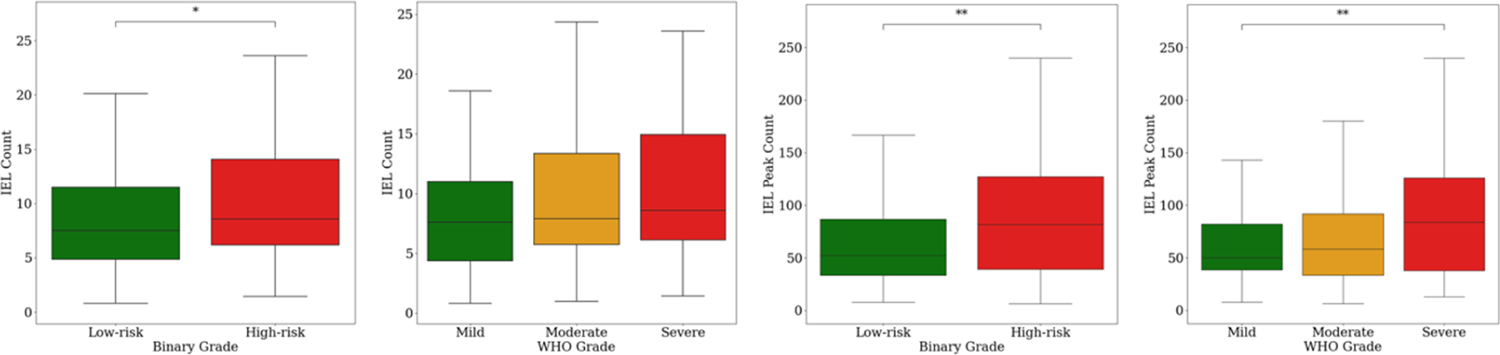
Boxplots showing the distribution of IEL scores (IC and IPC) in OED cases according to grade. For binary grade, low-risk cases are green and high-risk are red. For the WHO grade, mild cases are green, moderate orange, and severe are red.

Finally, we additionally visualise some of the results of this study, comparing cases that did transform with both high and low IC (and IPC) values, respectively, to a case that did not transform with a low IC (and IPC; see **Figure 5**). These images also show the hotspots used to generate the IPC scores. Overall, we see the hotspots tend to focus on the basal layer of the epithelium, which tend to have the highest density of IELs. Visibly, the case with higher IC/IPC appears to have many more IELs than the other cases with low scores.

**Figure 5.**
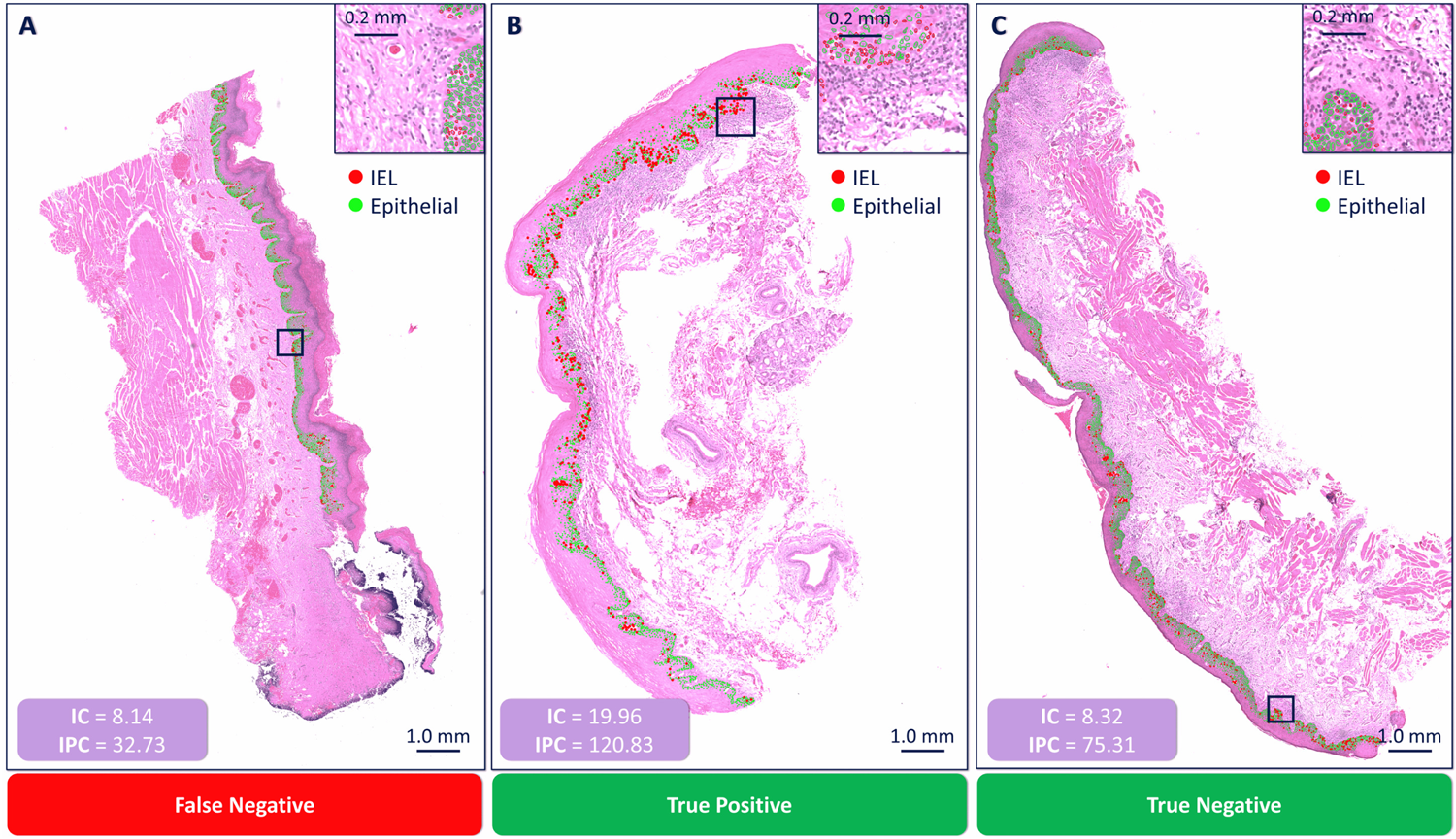
Visualisation of segmented nuclei within the dysplastic epithelium. Each panel shows a different WSI with the nuclear detections overlaid, where green dots are epithelial nuclei, and red dots are IELs. We additionally display the IC and IPC score for each slide, and the hotspot used to generate the IPC score. Panel A) shows a WSI that both the IC and IPC scores were low for, that did transform (i.e. a false negative). B) shows a WSI where the scores were high, and it transformed (i.e., a true positive); whilst C) shows a WSI where the scores were low, and it did not transform (i.e. a true negative). Within these analyses the cutoff values for the IC and IPC scores were 9.87 and 79.02, respectively.

## 4. Discussion

In this study, we investigated the prognostic potential of IELs derived from digital pathology in OED, a precursor lesion to OSCC. We highlighted the challenges associated with the current diagnostic and prognostic methods for OED, emphasising the need for more objective and reproducible approaches to predict the risk of malignant transformation. We addressed this gap by employing advanced deep learning techniques to automate the segmentation of dysplastic regions and IELs within oral tissue WSIs, and introduced four IEL scores as potential prognostic indicators, namely the II, IPI, IC, and IPC. Finally, we tested the prognostic utility of these scores in various survival models.

Our findings demonstrated the promising prognostic utility of the proposed digital IEL scores in predicting transformation-free survival in OED patients. Notably, the IEL scores generated based on the number of IELs per dysplastic epithelial cells (i.e., IC and IPC scores) showed the strongest association with clinical outcomes, as evidenced by their higher concordance indices and significant separation of low- and high-risk cases. The success of the count-based scores (i.e., IC and IPC) is perhaps unsurprising as they mimic the IEL scores used previously in the literature, typically seen in duodenal biopsies for studying coeliac disease [37,38]. We additionally suggest that the IC was more prognostic when compared to the IPC, as any spurious IEL detections by the deep learning models, could result in a substantially higher (or indeed lower) IEL score in any given patch, resulting in an incorrect hotspot region for calculating the IPC. By contrast, incorrect detections would be diluted in the IC. Comparison with traditional grading systems revealed that the binary grading scheme demonstrated clearer stratification of risk groups compared to the WHO grading system.

However, the inclusion of digital IEL scores provided additional prognostic information beyond both grading systems, as evidenced by the multivariate analyses. Furthermore, post-hoc analyses illustrated the significance of IEL scores in distinguishing between cases that progressed to malignancy and those that did not, with higher IEL scores consistently observed in transformed cases.

These findings suggest that incorporating digital IEL scores into existing grading systems could enhance their predictive accuracy and improve risk stratification in OED. Additionally, our study highlights the potential of computational pathology and deep learning techniques in identifying novel prognostic biomarkers and refining diagnostic and therapeutic strategies in head and neck cancer.

The potential positive association between inflammatory response and malignant transformation in OED is an interesting finding, challenging the conventional notion of immune cell infiltration being a favourable prognostic factor, which is often seen in cancer. However, we suggest that our finding may not be counterintuitive, with a higher immune response in dysplasia being indicative that a mechanism may already be underway (unseen on the simple H&E slide) in the action of transforming the OED lesion into cancer.

Furthermore, these results are consistent with previous studies that have observed a higher abundance of immune cells, including both IELs and PELs, in more dysplastic OED cases. Gannot *et al.* [39] noted increased immune cell infiltration in tongue lesions progressing to OSCC. Similarly, both Fitzpatrick *et al.* [40] and Hidalgo *et al.* [32] found a substantial number of OED cases to have band-like inflammatory cell infiltrate underlying the epithelium (i.e., PELs) and infiltrating the epithelium (i.e., IELs). However, the role of these immune cells is still subject to debate, with studies suggesting that severe dysplasia often become progressively infiltrated with immune suppressive myeloid cells and regulatory T cells (Treg), with Treg infiltration associated with an increased risk of malignant progression [41,42]. Yet, despite this, OED lesions are often strongly infiltrated with CD8+ T lymphocytes, that may act to reverse this immunosuppressive microenvironment [42,43]. Recent evidence has suggested a higher density of PELs in cases undergoing malignant transformation [26]. This was further supported by Shephard *et al.* [20], who additionally found a potential association between both PELs and IELs and malignant transformation.

The authors recognise some limitations regarding this study. Despite the work being based of a sizeable OED cohort (indeed one of the largest known digital OED cohorts to date), the dataset used was still relatively small. Moreover, all data was collected retrospectively from a single centre, and thus may be subject to certain biases. Further validation in larger, multicentric cohorts is warranted to confirm the generalisability and robustness of our findings. Additionally, the mechanistic underpinnings of the observed associations between IEL infiltration and malignant transformation in OED warrant further investigation.

In conclusion, our study contributes to the growing body of evidence supporting the role of immune cell infiltration as a potential prognostic indicator in OED. By elucidating the molecular mechanisms underlying the interactions between immune cells and dysplastic epithelial cells, we may uncover new avenues for personalized diagnostic and therapeutic strategies in head and neck oncology.

## Data Availability

We are unable to share the whole slide images and clinical data used in this study due to restrictions in the ethics applications.

## Acknowledgements

This study was supported by a Cancer Research UK Early Detection Project Grant, as part of the ANTICIPATE study (grant no. C63489/A29674) in addition to funding from the National Institute for Health Research (award no. NIHR300904). The authors express their gratitude to Professor Paul Speight (PMS), Professor Paula Farthing (PMF), Dr Daniel Brierley (DJB), and Professor Keith Hunter (KH) for their valuable contribution in providing the initial histological diagnosis.

## 6. Statement of Author Contributions

1. AS, HM, SAK and NMR designed the study with the help of all co-authors.
2. AS, HM and NMR developed the computational methods.
3. AS wrote the code and carried out the experiments.
4. SAK and HM obtained the ethical approval and retrieved the histological and clinical data from Sheffield.
5. All authors contributed to the writing of the manuscript.

